# Social connectedness as a determinant of mental health: A scoping review

**DOI:** 10.1101/2022.01.26.22269896

**Authors:** Priya J. Wickramaratne, Tenzin Yangchen, Lauren Lepow, Braja G. Patra, Benjamin Glicksburg, Ardesheer Talati, Prakash Adekkanattu, Euijung Ryu, Joanna M. Biernacka, Alexander Charney, J. John Mann, Jyotishman Pathak, Mark Olfson, Myrna M. Weissman

**Affiliations:** Department of Psychiatry, Vagelos College of Physicians and Surgeons, Columbia University Irving Medical Center, New York, NY, United States of America; Division of Translational Epidemiology, New York State Psychiatric Institute, New York, NY, United States of America; Departments of Psychiatry and Genetics & Genomic Sciences, Icahn School of Medicine at Mount Sinai, New York, NY, United States of America; Division of Health Informatics, Department of Population Health Sciences, Weill Cornell Medicine, New York, NY, United States of America; Department of Information Technologies and Services, Weill Cornell Medicine, New York, NY, USA; Department of Health Sciences Research, Mayo Clinic, Rochester, MN, United States of America; Division of Molecular Imaging and the Neuropathology, New York State Psychiatric Institute, Departments of Psychiatry and Radiology, Vagelos College of Physicians and Surgeons, Columbia University Irving Medical Center, New York, NY, United States of America

## Abstract

Public health and epidemiologic research have established that social connectedness promotes overall health. Yet there have been no recent reviews of findings from research examining social connectedness as a determinant of mental health. The goal of this review was to evaluate recent longitudinal research probing the effects of social connectedness on depression and anxiety symptoms and diagnoses in the general population. A scoping review was performed of PubMed and PsychInfo databases from January 2015 to December 2020 following PRISMA-ScR guidelines using a defined search strategy. The search yielded 56 articles representing 52 unique studies. In research with other than pregnant women, 84% (16 of 19) studies reported that social support benefited symptoms of depression with the remaining 16% (3 of 19) reporting minimal or no evidence that lower levels of social support predict depression at follow-up. In research with pregnant women, 80% (21 of 26 studies) found that low social support increased postpartum depressive symptoms. Among 3 of 4 studies that focused on loneliness, feeling lonely at baseline was related to adverse outcomes at follow-up including higher risks of major depressive disorder, depressive symptom severity, generalized anxiety disorder, and lower levels of physical activity. In 5 of 7 reports, smaller social network size predicted depressive symptoms or disorder at follow-up. In summary, most recent relevant longitudinal studies have demonstrated that social support protects adults in the general population from depressive symptoms and disorders. The results, which were largely consistent across settings, exposure measures, and populations, support efforts to improve clinical detection of high-risk patients, including adults with low social support and elevated loneliness.

## Introduction

While there is no universally accepted definition of social connectedness, it generally denotes a combination of interrelated constructs spanning social support, social networks, and absence of perceived social isolation. There is a broad-based agreement in the public health and epidemiologic literature that social connectedness protects and promotes mental and physical health and decreases all-cause mortality [1-3]. Researchers in fields ranging from psychology and epidemiology to sociology have been aware of these findings for several decades, but its implications have only recently begun to be appreciated more widely [4]. A recent study that reviewed the associations between social determinants of health and mental health outcomes [5], did not include social connectedness as a social determinant of mental health.

In a recent study of 100,000 participants in the UK Biobank [6], frequency of confiding in others and visits with family and friends emerged from over 100 modifiable risk factors as the strongest predictor of depression. This suggests that social connections may have protective effects or may be modified by development of a mood disorder. There is now a social connection domain in the Epic Social Determinants of Health wheel included in Electronic Health Records used in many major health organizations.

In light of these developments, we decided to conduct a scoping review of the relevant literature published between 2015 and 2020 to evaluate the extent to which social connection influences risk for depression and anxiety. We also sought to determine which aspects of social connections are most protective. Our review synthesizes recent literature on the various categories of social connection (social networks, perceived emotional support, and perceived social isolation) and their differential effects on depression and anxiety in specific populations. Because depression and anxiety can have adverse effects on social connections [7], we restricted our search to longitudinal/cohort studies from which appropriate temporal ordering that is necessary, although not sufficient, for causal inferences, can be established. We performed a scoping review of the literature published during the last five years addressing whether social connectedness is longitudinally associated with common mental health outcomes of depression and anxiety among adults.

## Methods

Searches of PubMed and PsychInfo databases and inspection of reference lists of relevant papers published during January 2015 and December 2020 were conducted following the Preferred Reporting Items for Systematic Reviews and Meta-Analyses Extension for Scoping Review (PRISMA-ScR) guidelines. The databases were searched using the following search strategy: (((“social* support *”or “social* isolation*” or “social* network*”) AND (Depression OR Anxiety)) AND ((“2015/1/1”[Date - Publication]: “2020/12/31”[Date - Publication]))) AND (English [Language]). Figure 1 presents a flow diagram displaying the process of searching and selecting the studies.

**Fig 1.**
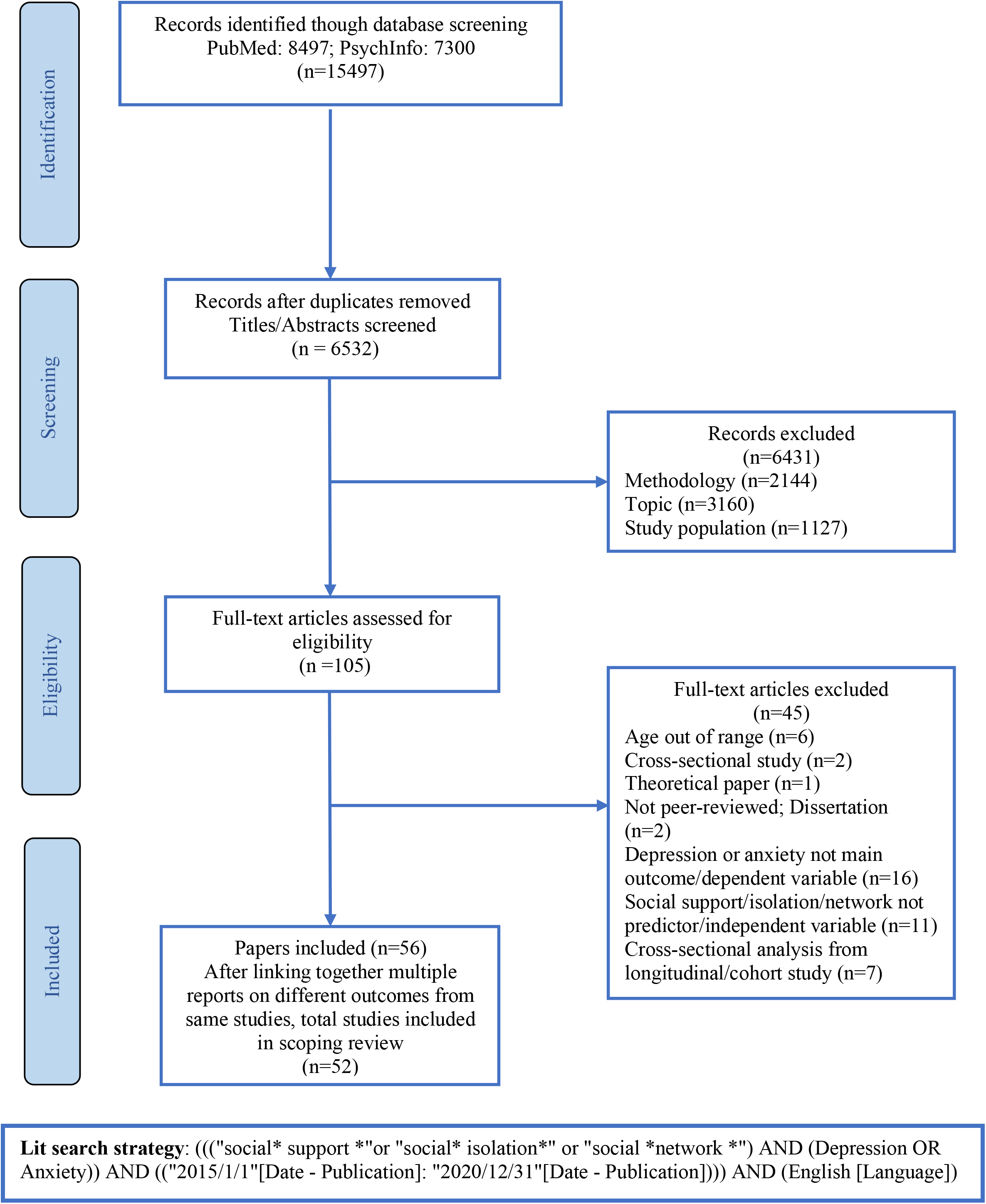
PRISMA flow chart of the scoping review. PRISMA diagram showing search and selection process of scoping review.

### Search strategy and selection criteria

For inclusion in the review, studies were required to meet the following criteria: (a) employed longitudinal/cohort study design; (b) published in peer-reviewed journals between the years 2015 and 2020 in English; (c) assessed social support, social networks, or social isolation as one of the main predictor variables; (d) the mental health outcomes analyzed in the articles had to be either depression or anxiety; and (e) recruited participants aged 18 years or older in the study.

Studies were excluded if: (a) the article did not report original data (e.g., the article was a theoretical paper, review paper, or meta-analysis); (b) social connectedness as operationalized in the review was not measured as a predictor variable; (c) the sample included participants with pre-existing health conditions (e.g., HIV, chronic disease, cancer, stroke, etc.) except mental health conditions that are generally comorbid with depression or anxiety; and (d) the study did not focus on adults.

### Data Extraction

After removing irrelevant titles and duplicates, the remaining articles were reviewed with respect to the eligibility criteria. Two authors (TY and PW) scrutinized titles and abstracts, and full text articles potentially eligible for this review were obtained. Key information from included papers was initially extracted and tabulated by one author (TY). The accuracy of this information was independently verified by another co-author (PW). Discrepancies were resolved through consensus. The data extracted included basic descriptive information about the sample, study type, length of follow-up, relevant predictor and outcome measures, and main findings. In cases where the same parent study data were employed in more than one publication, the papers were considered one study. Due to the wide variation in study designs and populations, we did not attempt meta-analysis, but rather provide a narrative synthesis of the main findings.

## Results

The initial search yielded 15,497 articles, and 6,532 articles were retained after duplicates and irrelevant articles were removed. After title and abstract screening, 105 articles were assessed for eligibility. Among studies selected for full-text review, 45 articles were deemed ineligible and excluded for various reasons: 10 did not employ longitudinal study design, 6 recruited participants aged younger than 18 years, 11 did not assess social support, social networks, and/or social isolation as predictor variable, 16 did not have depression or anxiety as the main outcome variable, and 2 were not from peer-reviewed sources. The PRISMA flowchart (Fig. 1) provides further detail on reasons for exclusion. Therefore, a total of 56 articles representing 52 unique studies met inclusion criteria for this scoping review.

## Study Characteristics

### Social Support

A little over half of the recent articles on the effect of social support on depression addressed the issues of depression in pregnant women, while the rest addressed the association of social support with anxiety or depression, in a variety of populations, other than pregnant women. We begin our review with nonpregnant populations because the results are of broader general relevance.

#### Study Samples other than pregnant women

Table 1a lists the 20 articles that reported quantitatively on the longitudinal effects of social support on depression or anxiety in samples other than pregnant women, but two papers reported on one study, resulting in a total of 19 studies. The sample size of the included studies ranged between 86 and 7171 participants. Majority of the studies were conducted in North America (8 studies), and the remaining were from Europe (6 studies), Asia (2 studies), and Australia (2 studies). One study was international in scope and sampled students from 76 host countries. All studies followed up the cohorts for at least one to two years and eight for over two years.

**Table 1a.**
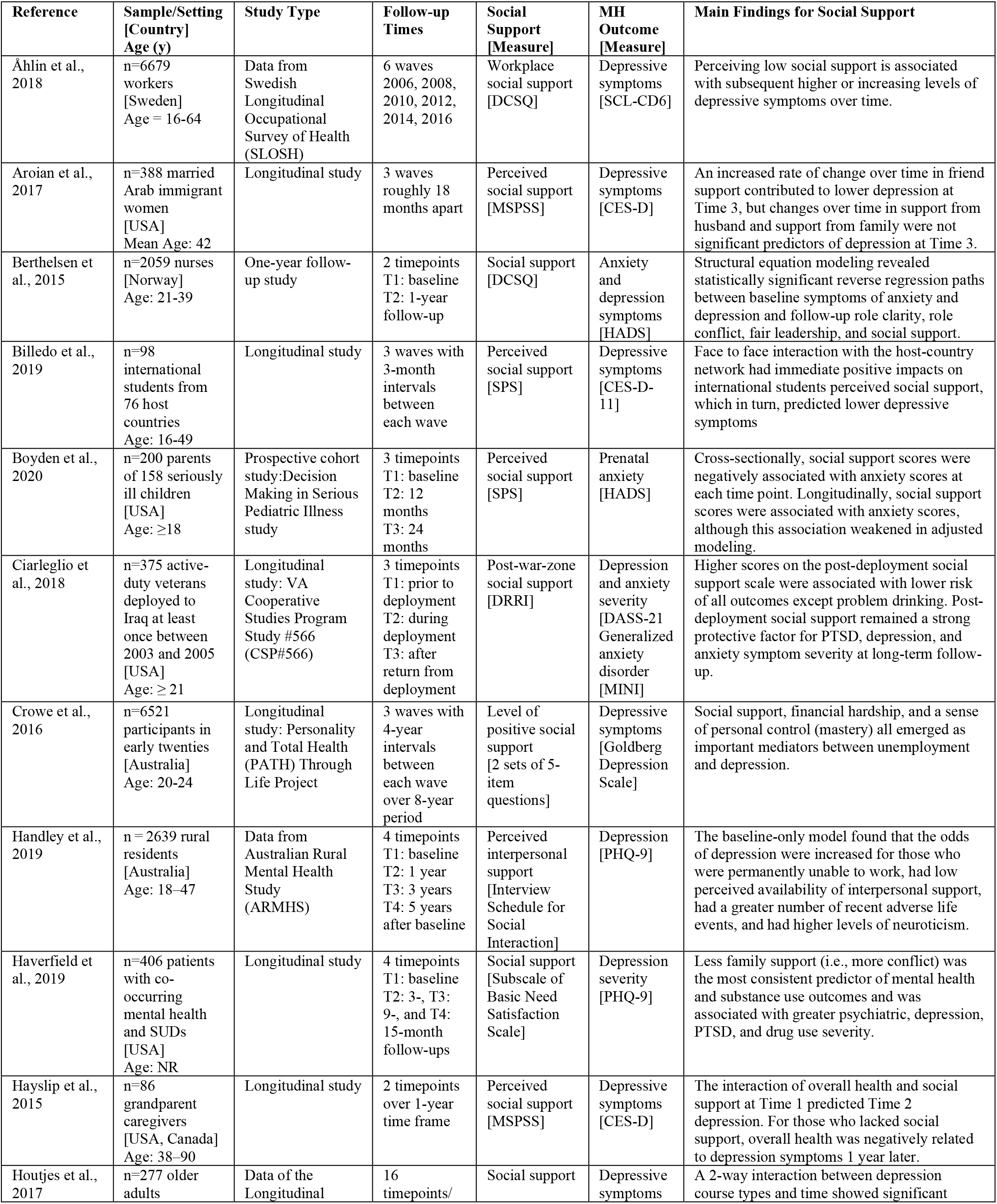
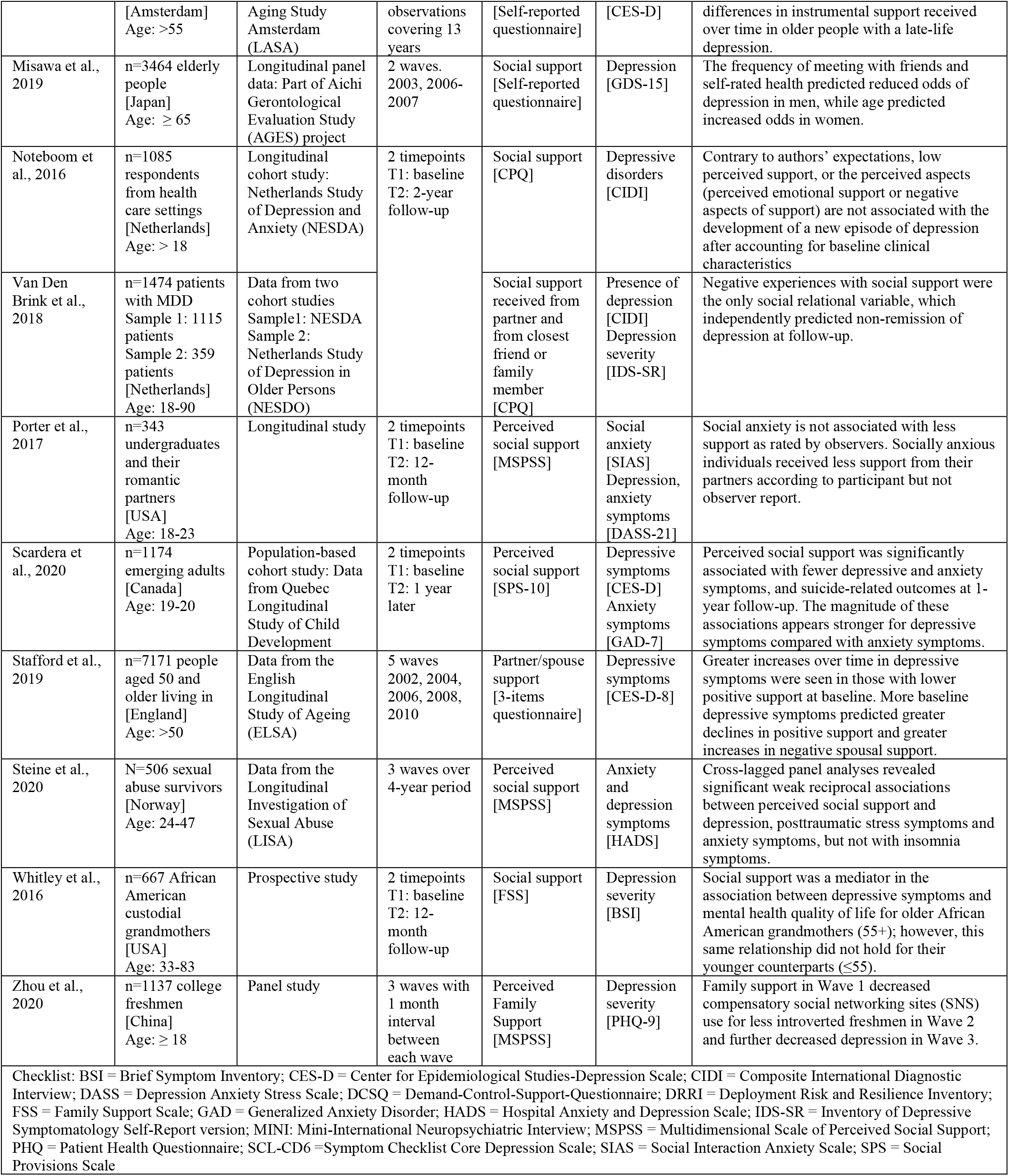
Characteristics of included studies on social support for nonpregnant samples.

#### Assessing Depression / Anxiety and Social Support

The nineteen included studies used a threshold score on a depression rating scale to measure depressive symptoms, and the Center for Epidemiological Studies Depression Scale was the most frequently utilized scale, being used in six of the studies. Other measures with established psychometric properties included in this Table were as follows: Patient Health Questionnaire-9, Geriatric Depression Scale, Inventory of Depressive Symptomatology, Symptom Checklist Core Depression Scale, and Brief Symptom Inventory. In order to assess anxiety and depressive symptoms, three studies [8,9,10] used the Hospital Anxiety and Depression Scale, and two [11,12] used Depression Anxiety Stress Scale. Additionally, one study [13,14] used Composite International Diagnostic Interview (CIDI), a structured diagnostic interview to measure the presence of a major depressive disorder (MDD) according to DSM-IV criteria. The social support measures in these studies varied considerably, with five studies [10,12,15-17] using the original or the adapted version of the Multidimensional Scale of Perceived Social Support (MSPSS). Two studies [8,18] employed Demand-Control-Support-Questionnaire (DCSQ) to assess workplace social support, and three studies [9,19, 20] used Social Provision Scale (SPS). A scale in the Deployment Risk and Resilience Inventory (DRRI) was employed in one veteran study to assess the extent to which they perceive assistance and encouragement in the war zone from fellow unit members [11]. Four studies [21-24] used questionnaires that were developed by the authors. The Interview Schedule for Social Interaction—availability of Attachment Scale was used to measure the perceived availability of interpersonal support [25]. Whitley et al [26] used the Family Support Scale (FSS), and Haverfield et al. [27] used a subscale of the Basic Need Satisfaction Scale to assess general social support (Table 1a).

#### Effects on Depression/Anxiety

Social support has been shown benefit in abating symptoms of depression over time in 16/19 or 84% of the studies (Table 1a). Analyses of mental health conditions in a sample of nationally dispersed war-zone veterans for over seven years indicated that higher levels of social support post-deployment were associated with decreased risk of depression and anxiety disorders, as well as less severe symptoms [11]. Moreover, reduced levels of perceived support at the workplace were associated with increased levels of depression symptoms [18], which aligns with previous research that showed a direct influence of social support on the well-being of medical staff workers in the subsequent study waves [8]. A five-year study of rural community residents also found that low perceived interpersonal support was associated with adverse mental health outcomes, including depression [25]. Billedo et al. [19] found short-term reciprocal associations between social support and depressive symptoms in sojourning students. They further stated that face-to-face interaction with the host-country network had immediate positive effects on perceived social support, which subsequently predicted lower depressive symptoms [19]. Even in samples of emerging adults, higher levels of perceived social support were protective against depressive and anxiety symptoms [20,21]. Boyden and colleagues followed a sample of parents of critically ill children over two years and found that greater perceived social support was associated with lower anxiety levels across assessments [9]. Higher baseline social support remained negatively associated with lower parental anxiety scores at 12 months (B=-0.12, *p*=0.03; 95% CI =-.23 to -.01) and 24 months (B=-0.11, *p*=0.04; 95% CI=-0.21 to -0.01). This inverse association had dissipated by 24 months in their adjusted modeling [9]. Their findings concur with other aforementioned studies that demonstrated a benefit of having supportive relationships.

Interestingly, in terms of the source of social support, the role of family support remains unclear. One study of married Arab immigrant women in the U.S. did not find family support to be protective against depression. Instead, support from friends was found predictive of fewer depressive symptoms at follow-up [15]. However, the results of this study contrast with those of Zhou et al., [17] who showed that more perceived family support decreased usage of social networking sites, which was followed by decreased depressive symptoms among Chinese college students. Similarly, Haverfield et al. [27], analyzed data from patients with co-occurring disorders at treatment intake and across follow-ups in the United States and found that deficits in family support were the most consistent predictor of greater depression and substance use severity [27]. Consistent with previous studies on social support drawn from family and their positive impacts, two studies [16,26] specifically focused on custodial grandparents in the U.S. showed that while elevated caregiving stress may negatively affect grandparent caregivers’ mental and overall health over time, greater social support from family networks may reduce depression accompanying caregiving. According to Whitley et al., [26] social support has a mediating effect on the relationship between depression and mental health quality of life in older (55+) African American custodial grandmothers, but not in their younger (≤55) counterparts.

Although depression peaks in young adulthood, it either can persist or emerge later in life, as evidenced by the three studies focusing exclusively on the role of social support in samples of adults aged 55 and older. Misawa and Kondo (2019), in a study of 3464 Japanese older people, reported that social support was unrelated to changes in depressive symptoms but added that social factors of having hobbies and meeting frequently with friends were associated with improved late-life depression. This link between social interaction and social support was only observed to be protective for men [23]. Conversely, another study spanning eight years of follow-up of older adults in England found a bidirectional association between depressive symptoms and spousal support [24]. The authors found an average decrease in positive support (or an increase in negative support) over time in age- and gender-adjusted models, which later predicted increasing depressive symptom trajectory [24] Another study on older people with depression revealed that a chronic course of depression might decrease received social support over time [22]. Moreover, their findings suggest that pre-existing depression in concert with less social support may predispose older persons, especially men and single people, to more depression over time [22].

A few studies (3/19 = 16%) found minimal or no evidence that lower levels of social support predict depression at follow-up. For example, in a naturalistic cohort study of Noteboom et al. [13] people with a prior history of depression reported a smaller network size and less emotional support at baseline However, these structural (network size or having a partner) and the perceived aspects of social support had no predictive value in the longitudinal data. Similarly, only negative experiences with social support proved to be a risk factor for non-remission, independent of other social-relational variables in depressed persons [14]. Steine et al. [10] found statistically significant weak reciprocal associations between perceived social support and depression and anxiety symptoms over time among adult survivors of childhood sexual abuse. Although, Porter and Chambless reported a link between higher odds of relationship dissolution and social anxiety, they observed no differences between participants with high versus low social anxiety with regard to the amount of social support provided by their partners [12].

#### Social support for women during and post pregnancy

Table 1b presents results from twenty-eight articles examining associations between social support and depression during pregnancy or postpartum, uniquely vulnerable periods for women during which they may experience a range of psychosocial stressors. Two pairs of papers provided findings based on identical samples and were reported in a combined table entry, resulting in twenty-six studies. Studies were predominantly conducted in Asia (42%) and North America (31%), with the remaining studies from Europe (15%), South America (4%), Africa (4%), and Australia (4%). In terms of individual countries, four studies were conducted in the United States, followed by Canada, China, Japan, and Turkey, all with three studies each. Sample sizes varied from 54 participants in a follow-up of a previous randomized trial [28] to 12,386 couples in a study with findings on both maternal and paternal depression [29]. Most of the included studies [28,30-43] began in the mid or late pregnancy, three [44-46] started in early pregnancy, and eight studies [29,47-53] started after birth. Women were on average aged between 18 and 40 years old. The most commonly used measures of social support and depression were, respectively, the Medical Outcomes Study Social Support Survey (MOS-SSS) and the Edinburgh Postnatal Depression Scale (EPDS) (Table 1b).

**Table 1b.**
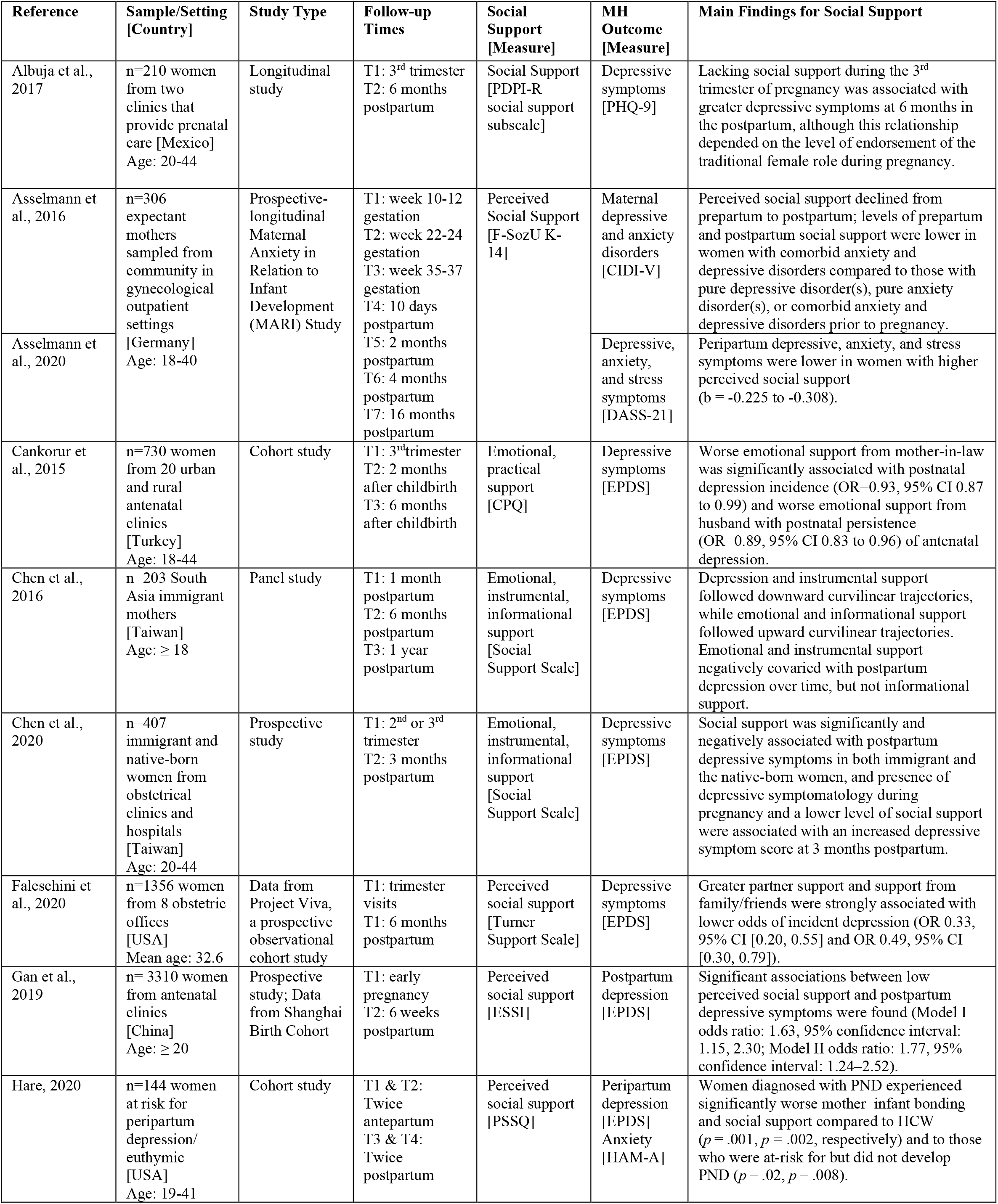
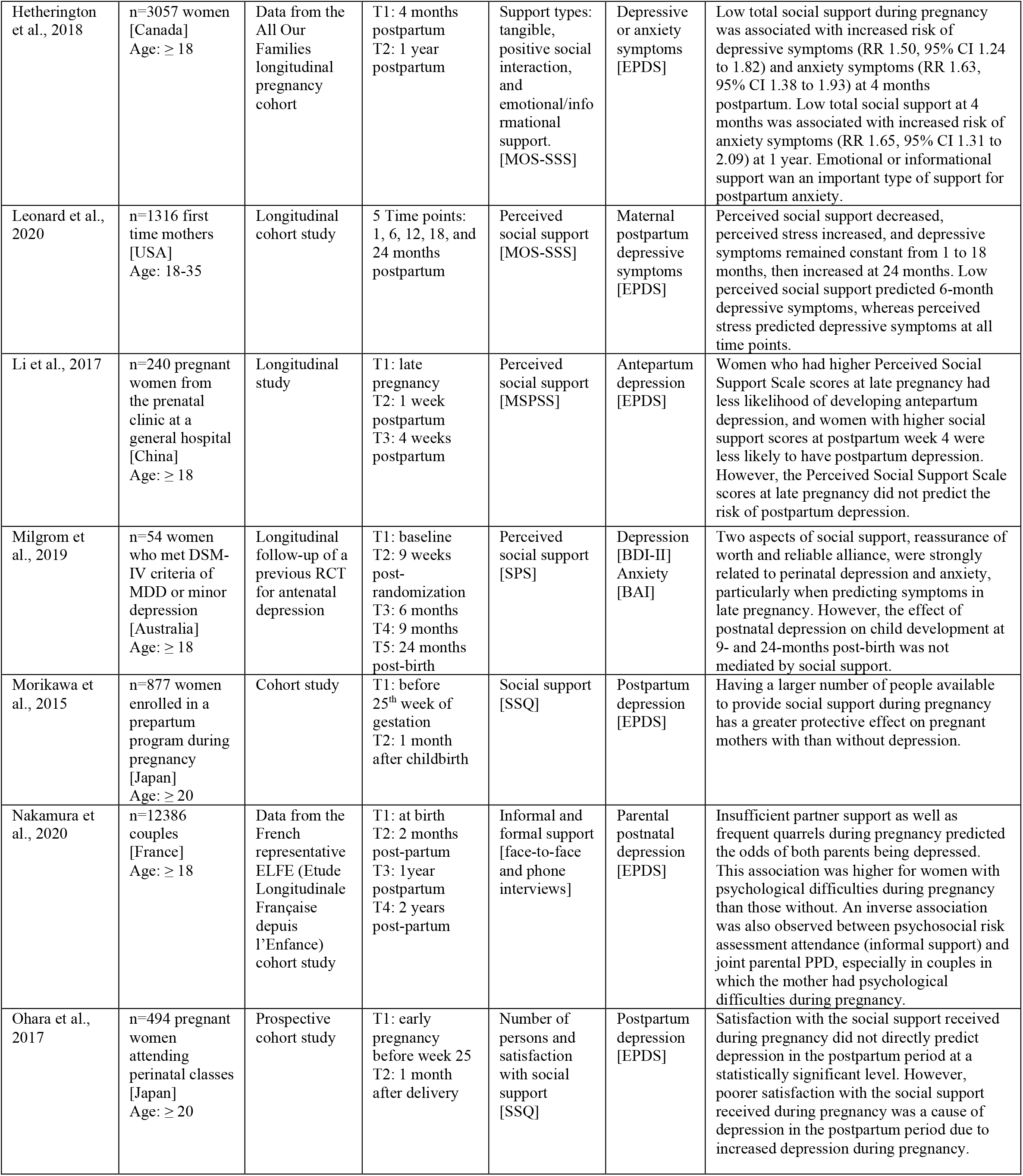
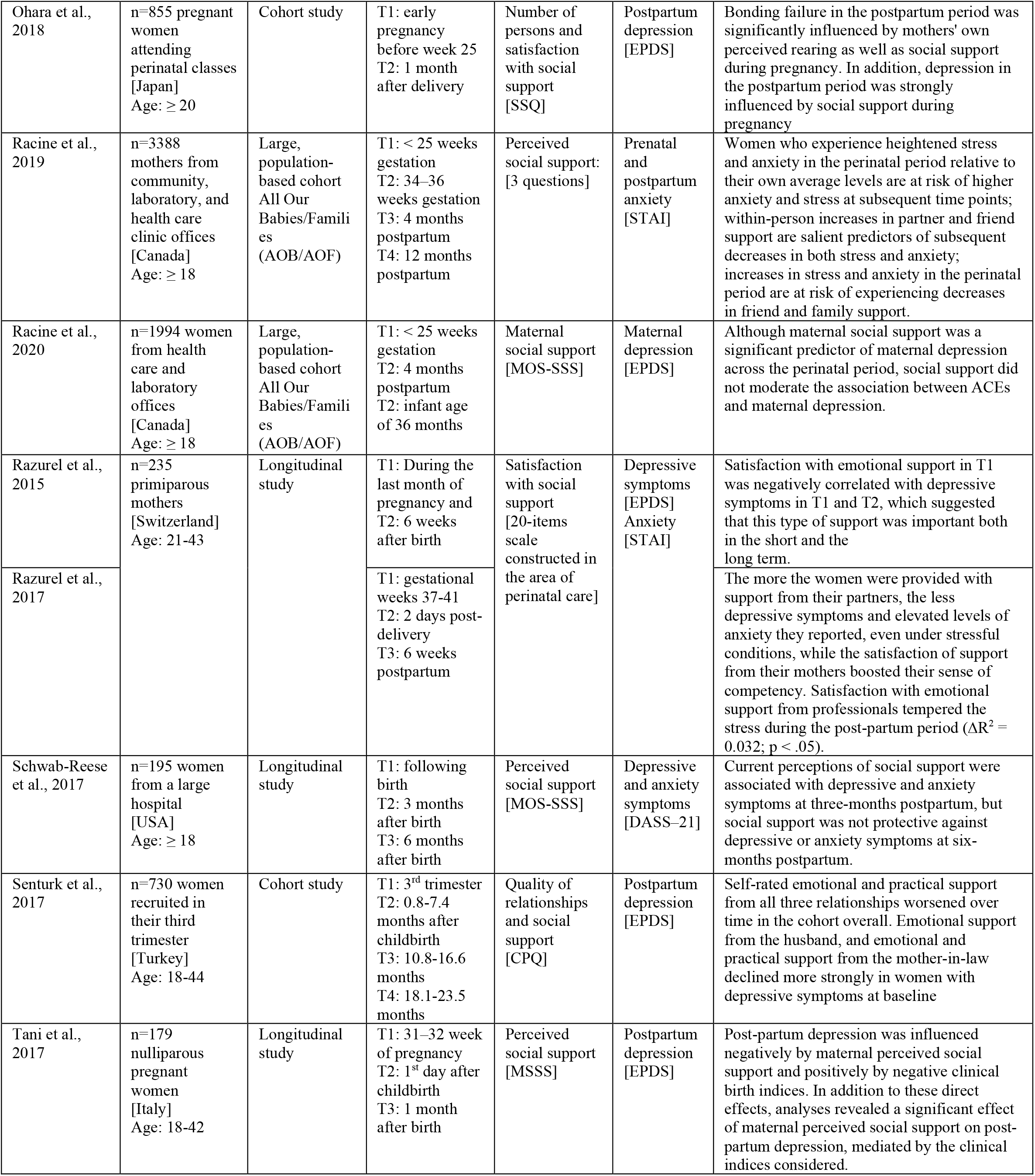
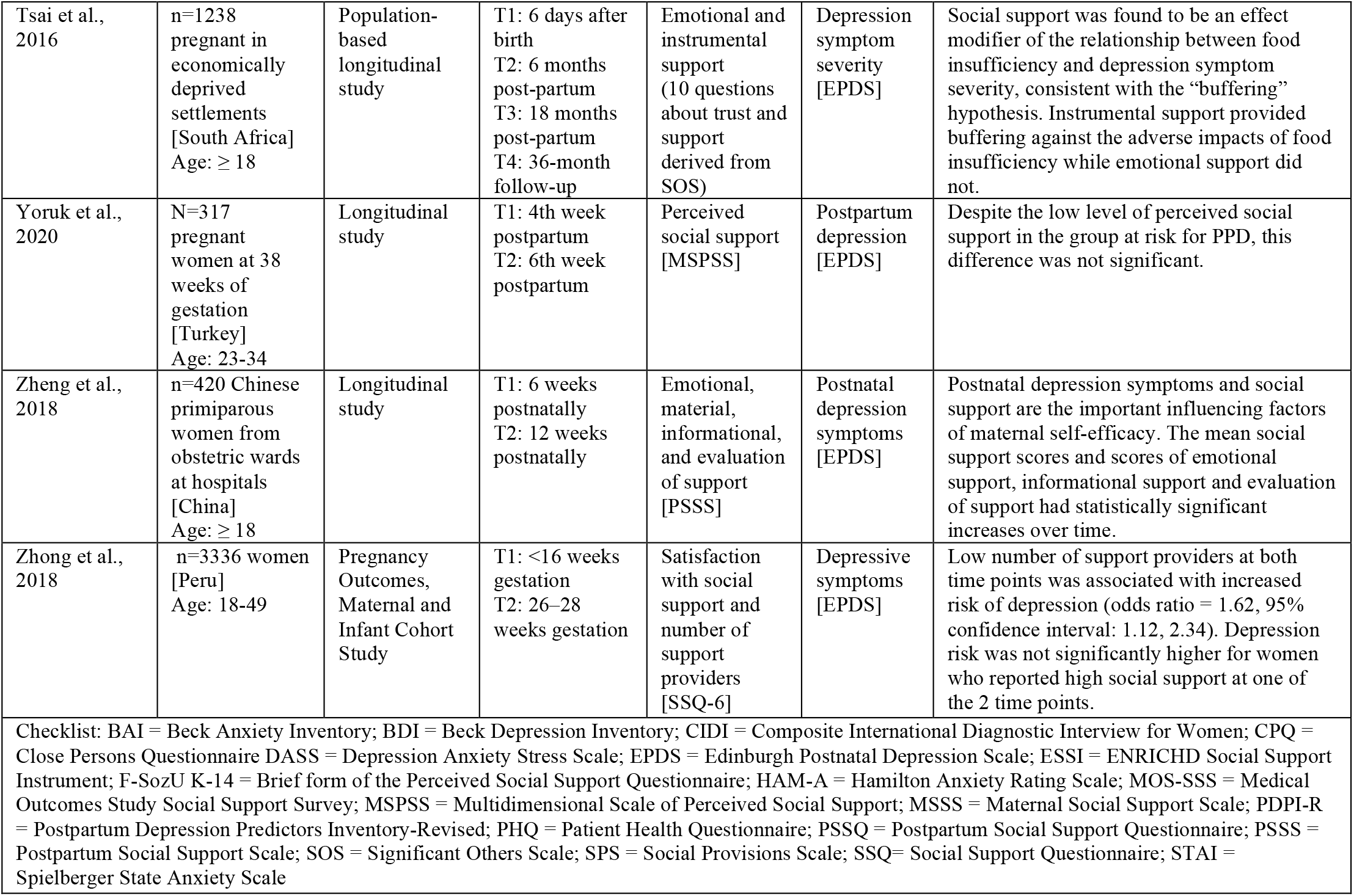
Characteristics of included studies on social support for pregnant women.

The majority of the studies (21/26 = 80%) found low social support increased postpartum depressive symptoms. Women with higher perceived social support exhibited lower depression (b = -0.308, SE = 0.036, p < .001) and anxiety (b = -0.225, SE = 0.039, *p* < .001) symptom severity across the peripartum period [31]. Similar results were obtained by other studies [28,38,43,44,48] conducted to investigate the trajectory of the association between depression and perceived social support. A study in Taiwan comprising 407 immigrant and native-born women showed that a presence of depression symptoms during pregnancy (β = 0.246; *p*<0.001) and deficient social support (β = -0.233; p<0.001) positively covaried with depressive symptoms at 3-months postpartum [34]. The significant protective factor of social support against postpartum depression was also highlighted in another study through a mediating effect of good clinical delivery.^46^ Moreover, insufficient social support and frequent quarrels during pregnancy were associated with significant increase in joint postpartum depressive symptoms in mothers and fathers [29]. Tani et al. demonstrated a protective role of maternal and paternal relationships on postpartum depression in nulliparous women [42].

Asselmann and colleagues [32] found a bidirectional relationship between peripartum social support and psychopathology. They concluded that low social support increased the risk for anxiety and depressive disorders, and these disorders before pregnancy, in turn, fostered dysfunctional relationships and lowered social support across the peripartum period [32]. Women with comorbid anxiety and depression were at higher risk for lacking social support during this timeframe [32]. Social support appeared to be a predictor of depression from mid-pregnancy to six months postpartum, particularly in late pregnancy. Nonetheless, the predictive effect of social support on anxiety was only observed in late pregnancy [43]. Latina women who had lower social support during the third trimester of pregnancy were reported to be at greater risk of depressive symptoms at six months in the postpartum, which was consistent with a study on primiparous mothers [49]. Specifically, we found that women with lower social support and higher self-reported adherence to the Traditional Female Role were at the highest risk of experiencing postpartum depressive symptoms [30].

Data from Canada reported inverse associations between 1) social support during pregnancy and anxiety and depression postnatally (RR 1.50, 95% CI 1.24 to 1.82) and 2) social support at four months postpartum and one year postpartum (RR 1.65, 95% CI 1.31 to 2.09) [48]. In contrast to these studies, Schwab-Reese et al. stated that social support was not protective against depressive or anxiety symptoms at six months postpartum as it was at three months postpartum [50].

Satisfaction with all the types of support (emotional, material, esteem, and informative) from the spouse reduced the psychological disorders in mothers as much in the prenatal compared to the postpartum period [40]. Among immigrant mothers in Taiwan from China or Vietnam, emotional support was found to be significantly and inversely associated with postpartum depression [34]. Emotional and informational support were identified as the most important types of social support for postpartum anxiety [48]. Marginal structural models were employed to evaluate the time-varying associations of low social support (before and in early pregnancy) with depression in late pregnancy in a cohort of Peruvian women, and analyses suggested that women with sustained low scores on the 6-item Sarason Social Support Questionnaire (SSQ-6) were at higher risk of antepartum depression [43]. The authors also found a stronger association of fewer persons providing social support on depression risk than low social support satisfaction.

Others have found that not all forms of social support during a woman’s transition into motherhood are equally beneficial in alleviating depression and anxiety symptoms. Razurel et al. reported that support provided by one’s partner buffered the effects of stress on depression, although support from friends or professionals did not [39]. Although social support from family and friends was deemed less prominent than that from the spouse, studies [35,40] showed that these sources of support also had an influence on maternal mental health. A study evaluating the impact of family relationships and support on perinatal depressive symptoms between the third trimester of pregnancy and two to six months postpartum noted that the incidence and persistence of depression symptoms were predicted by lower baseline perceived emotional support from the mother-in-law and the husband, respectively [33]. In a large population-based study, increases in partner and family support had a more protective effect against anxiety and stress, and higher than average levels of anxiety and stress led to maternal-reported decreases in support [54].

Some studies [37,52] did not find associations between low perceived social support during pregnancy and postpartum depression. Ohara et al. [46] showed that satisfaction with social support did not directly predict depression in the postpartum period at a statistically significant level. Further, their path model revealed that less satisfaction with the social support received during pregnancy was rather a cause of postpartum depression. This indirect link of social support with depression in the postpartum period is in line with another study with slightly larger sample size, one year later, by the same first author [45]. While one study [55] found that the number of supportive persons during pregnancy had a more substantial effect on decreasing postpartum depressive symptoms in depressed relative to non-depressed mothers, their analyses also showed that satisfaction with social support was not a significant predictor of postpartum depression. The inconsistent findings on social support and depression and anxiety across included studies can be partially attributed to varying operational definitions of social support, utilization of different self-report social support and depression measures, and insufficient control for confounding variables.

### Social isolation/Loneliness

Given that both these concepts were concurrently assessed in addressing mental health outcomes in some studies, articles that met the inclusion criteria of the current review and examined the subjective counterpart of social isolation were included in Table 2. This table presents characteristics of selected studies that explored the impact of social isolation on depression and anxiety disorders. The four studies [56-59] reported on a total of 7,820 older adults aged 50 and above in European countries, namely Ireland, Wales, Germany, and the Netherlands. The number of participants in the studies varied considerably, from 285 participants with a primary diagnosis of depression in a two-year follow-up study to 5,066 in a report on a well-characterized cohort of adults from Ireland [56]. Social isolation and loneliness (perceived social isolation) are intricately related, albeit conceptually distinct.

**Table 2.** Characteristics of included studies on social isolation.

| Reference | Sample/Setting [Country] | Study Type | Follow-up Times | Social Isolation [Measure] | MH Outcome [Measure] | Main Findings for Social Isolation |
| --- | --- | --- | --- | --- | --- | --- |
| Domènech-Abella et al., 2019 | n=5066 adults [Ireland] age ≥50 | Irish Longitudinal Study on Ageing (TILDA) | 2 waves of TILDA<br>Second wave: 2012-13<br>Third wave: 2014-15 | UCLA Loneliness Scale | Major depressive disorder (MDD) or generalized anxiety disorder (GAD) [CIDI] | The longitudinal association between experiencing loneliness and higher likelihood of suffering from GAD two years later is bidirectional, whereas the association between social isolation and higher likelihood of subsequent MDD or GAD as well as those between loneliness and subsequent MDD or deterioration of social integration are unidirectional. |
| Evans et al., 2019 | n=2135 elderly residents [Wales] Age: ≥65 | Data from the Cognitive Function and Ageing Study–Wales (CFAS–Wales) | 2 timepoints<br>T1: baseline<br>T2: 2-year follow-up | LSNS-6 | Depression and anxiety [AGECAT] | Older people with depression or anxiety perceived themselves as more isolated than those without depression or anxiety, despite having an equivalent level of social contact with friends and family. In people with depression or anxiety, social isolation was associated with poor cognitive function at baseline, but not with cognitive change at 2-year follow-up. |
| Herbolsheimer et al., 2018 | n=334 community-dwelling older adults [Germany] Age: 65-84 | Longitudinal study | 2 timepoints<br>T1: baseline<br>T2: 3 years later | LSNS-6 | Depressive symptoms [HADS] | Being socially isolated was associated with lower levels of out-of-home physical activity, and this predicted more depressive symptoms after 3 years. However, no direct relationship was observed between social isolation from friends and neighbors at the baseline and depressive symptoms 3 years later. |
| Holvast et al., 2015 | n=285 older adults [Netherlands] Age: ≥ 60 | Multi-site prospective cohort study from the Netherlands Study of Depression in Older Persons (NESDO) | 2 timepoints<br>T1: baseline<br>T2: 2-year follow-up | De Jong Gierveld Loneliness scale | Depression [CIDI]<br>Depression severity [IDS-SR] | Loneliness, subjective appraisal of social isolation, was a significant positive determinant of depressive symptom severity during follow-up. This association was independent of social network size and persisted after controlling for other potential confounders. |
| Checklist: AGECAT = Automated Geriatric Examination; CIDI = Composite International Diagnostic Interview; HADS = Hospital Anxiety and Depression Scale; IDS-SR = Inventory of Depressive Symptomatology Self-Report version; LSNS = Lubben Social Network Scale |  |  |  |  |  |  |

Domènech-Abella and colleagues (2019) found social isolation and loneliness to be antecedent risk factors for incident depression or exacerbating late-life generalized anxiety or major depressive disorder independently [56]. The authors cautioned readers not to underestimate the subjective aspects of social isolation. Their findings are concordant with a previous study by Holvast et al. [59] which showed that loneliness was independently associated with more severe depressive symptoms at follow-up (*β* = 0.61; 95% CI 0.12–1.11). Moreover, participants who scored high on the subjective appraisal of social isolation, measured by six-item de Jong Gierveld Loneliness Scale with subscales for emotional loneliness (perceived absence of intimate relationships) and social loneliness (perceived lack of a wider circle of friends and acquaintances), at baseline had a lower likelihood of achieving remission two years later [59]. The Composite International Diagnostic Interview (CIDI) was employed in both studies to measure depressive disorders at baseline [56,59], and Inventory of Depressive Symptoms (IDS) was used to assess the depression course at the follow-up in one study [59].

Two studies [57,58] utilized the Lubben Social Network Scale, a validated instrument designed to gauge social isolation in the elderly. Analysis of Cognitive Function and Ageing Study-Wales (CFAS-Wales) by Evans et al. (2019) showed that people with depression or anxiety experience poorer social relationships and higher social isolation and loneliness relative to those without such symptomatology, despite reporting an equivalent level of social contact [57]. The apparent reductions in negative affect at follow-up in over half of the respondents with clinically relevant depression or anxiety, diagnosed using the Automated Geriatric Examination for Computer-Assisted Taxonomy (AGECAT) algorithm, at baseline could be due to biopsychosocial changes intrinsic to ageing and/or cohort effects. Results by Herbolsheimer et al. (2018) opposed previous findings, stating that social isolation from friends and neighbors at the baseline was not directly associated with depressive symptoms at the 3-year follow-up [58]. Of importance, the authors also showed that being socially isolated from friends and neighbors was related to lower levels of out-of-home physical activity, which predicted more depressive symptoms after three years (*β* = .014, 95% CI .002 to .039) [58]. These reported findings demonstrate the potentially detrimental effect of objective and subjective social isolation on mood disorders in later life. Extant evidence suggests the importance of considering both social isolation and loneliness without the exclusion of the other in efforts to mitigate risk.

### Social networks

Five studies, i.e., seven papers, were identified that examined the longitudinal relationship between social networks and depression or anxiety (see Table 3). Four [56,60-63] of the five studies [13,14,56,60-63] on social networks focused on older adults. Community-dwelling individuals aged 75 and older with restricted social networks were more likely to develop depression compared to those who maintained an integrated social network [61]. While it has also been noted that respondents who experienced social loss within the last six months reported a higher risk of depression in old age, the adverse effects of loss on depression could be attenuated by the existence of an integrated social network [61]. In a large follow-up study spanning ten years that recruited older female nurses, lower social networks increased the risk of incident late-life depression in age-adjusted models [60]. In line with these results, another study on the elderly showed that a smaller network size measured using the Berkman-Syme Social Network Index is a robust risk factor of major depression and generalized anxiety disorder [56]. Conversely, one study on American adults aged 57–85 years denies this relationship as no direct effects of social networks on frequencies of depression symptoms were detected at follow-up [63]. It is worth noting that the same study also showed that social networks predicted higher levels of perceived isolation (β=0·09; p<0·0001), which in turn predicted higher levels of depression and anxiety symptoms (β=0·12; p<0·0001) [63]. According to papers [13,14] that used data from NESDA and NESDO, the size of social networks at baseline did not predict depression at follow-up. For adults with a pre-existing diagnosis of major depressive disorder, only the social network characteristic of living in a larger household was reported to have a unique predictive value for depression course [14].

**Table 3.** Characteristics of included studies on social network.

| Reference | Sample/Setting [Country] | Study Type | Follow-up Times | Social Network [Measure] | MH Outcome [Measure] | Main Findings for Social Network |
| --- | --- | --- | --- | --- | --- | --- |
| Chang et al., 2016 | n=21728 elderly women [USA]<br>age: ≥65 | Prospective cohort study: Nurses' Health Study (NHS) | 5 timepoints, Baseline and 4 biennial follow-up questionnaire cycles over 10-year period | Berkman-Syme Social Network Index | Depression [MHI-5 subscale of the SF-36, CESD-10, GDS-15] | Social factors (lower social network; lower subjective social status; high caregiving burden to disabled/ill relatives) were associated with higher incident late-life depression risk in age-adjusted models. |
| Domènech-Abella et al., 2019 | n=5066 adults [Ireland]<br>age ≥50 | Irish Longitudinal Study on Ageing (TILDA) | 2 waves of TILDA<br>Second wave: 2012-13<br>Third wave: 2014-15 | Berkman-Syme Social Network Index | MDD or GAD [CIDI] | Both objective social isolation (size of social network) and loneliness factors have been found to be robust risk factors for depression and anxiety independently, which acts as a warning not to underestimate the subjective aspects of social isolation. |
| Förster et al., 2018 | n=783 elderly people [Germany]<br>Age: ≥ 75 | Population-based cohort study: Leipzig Longitudinal Study of the Aged (LEILA) | 3 timepoints<br>T1: baseline<br>T2: follow-up1<br>T3: follow-up2 | PANT | Depressive symptoms [CES-D] | Persons with a restricted social network were more likely to develop depression. Risk of depression was particularly high for elderly with social loss experiences. An integrated social network may buffer the negative effects of loss on depression. |
| Noteboom et al., 2016 | n=1085 respondents from health care settings [Netherlands]<br>Age: > 18 | Longitudinal cohort study: Netherlands Study of Depression and Anxiety (NESDA) | 2 timepoints<br>T1: baseline<br>T2: 2-year follow-up | Questions on how many relatives, friends or others over the age of 18 years they had regular and important contact | Depressive disorders [CIDI] | Structural (network size and partner status) did not predict depression at follow up. The group with a lifetime history of depression reported a smaller social network. |
| Van Den Brink et al., 2018 | n=1474 patients with a major depressive disorder [Netherlands]<br>Age: 18 - 90 | Data from 2 cohort studies NESDA and Netherlands Study of Depression in Older Persons (NESDO) |  | Questions on social network characteristics | Presence of depression [CIDI]<br>Depression severity [IDS-SR] | Social network characteristics, such as having a partner and number of persons in one's household, are related to depression course. |
| Reynolds et al., 2020 | N=3005 elderly people [USA]<br>Age: 57–85 | Panel data from National Social Life, Health, and Aging Project (NSHAP) | 3 waves 2005, 2010, and 2015 with 5-year intervals between each wave | Questions on community-layer, interpersonal-layer, and partner-layer connection | Depressive symptoms [CES-D] | Results demonstrate multiple links between social connection and depression, and that the evolution of social networks in older adults is complex, with distinct mechanisms leading to positive and negative outcomes. Specifically, community involvement showed consistent benefits in reducing depression. |
| Santini et al., 2020 |  |  |  | Social Disconnectedness Scale | Depressive symptoms [CES-D-ML]<br>Anxiety [HADS-A] | No evidence that social disconnectedness (ie, having a small network or infrequent social interaction) predicted higher frequencies of depression symptoms at subsequent timepoints (β=0.003; p=0.77; data not shown) was found. Social disconnectedness predicted higher amounts of perceived isolation, which in turn predicted higher amounts of depression and anxiety symptoms. |
| Checklist: CES-D = Center for Epidemiological Studies-Depression Scale; CES-D-ML = Center for Epidemiological Studies-Depression Minus Loneliness Scale; CIDI = Composite International Diagnostic Interview; GAD = Generalized Anxiety Disorder; GDS = Geriatric Depression Scale; HADS = Hospital Anxiety and Depression Scale; IDS-SR = Inventory of Depressive Symptomatology Self-Report version; MDD = Major Depressive Disorder; MHI-5 = Mental Health Index-5 |  |  |  |  |  |  |

While smaller social networks emerged as a predictor of depression in three of the five studies, the remaining studies showed no predictive value of social networks in the depression course (2/5 = 40%). The extent to which measures assessed the concept of social networks across the included studies could be disputed and may account for between-study inconsistencies. Some assessed using a count of the number of people in the social network [13,14], one assessed social network type (restricted vs. integrated) [61], four assessed network size and frequency of interaction with network members [56,60,62,63], and one study assessed additional indicators such as three layers of human relationships (community-layer, interpersonal-layer, and partner-layer connection) [62]. These longitudinal studies found that social networks have both direct and buffering effects on psychological wellbeing.

## Discussion

The goal of this scoping review was to determine current knowledge regarding the effect of social connection on depression and/or anxiety symptoms and diagnoses in the general population. Lund et al. (2018) noted that while there is no universal definition of social connection, it is considered an umbrella term that generally includes the following components: social support, social networks, social isolation and loneliness [64].

Much of the research investigating the role of social connections in the studies published in the last five years has focused primarily on social support as compared to the other components of social connection and, more specifically, on perceived emotional support. The majority of studies, regardless of whether the support was received or perceived, showed that, in general, social support was a protective factor for both depressive symptoms and disorders. The consistency of the findings across a variety of settings, measures of exposure, and populations supports the generalizability of the findings in this review. The measures used varied in the type of social support assessed, although most studies assessed subjective and objective emotional support rather than structural support.

Few of the studies examined the effect of social support on anxiety symptoms or diagnoses, and those that did investigate these associations found the results mixed and weak. A single study examined the effect of social support on social anxiety and found no association [12]. However, there were too few studies to draw any firm conclusions.

Over half of these studies investigated the effects of social support on depression/ anxiety at various stages of pregnancy. The preponderance of studies that investigated the effect of social support on pre/postpartum depression could be due to our restriction of included studies in this review to longitudinal studies since the effects of social support during pregnancy on postpartum depression can readily be assessed by employing longitudinal study design.

There were far fewer studies that examined the longitudinal relationship between social networks and anxiety or depression, the majority of which focused on older adults. Although measures which assessed the concept of social networks were somewhat inconsistent and may have led to mixed results between studies, on the whole, social networks have direct and buffering effects on depression /anxiety. More studies of the effects of social networks such as size and structure and the characteristics of network ties, frequency of contact, reciprocity, duration, and intimacy will give us a greater understanding by which these networks impact depression and anxiety, and opportunities for intervention.

A small number of studies met our inclusion criteria for exploring the impact of social isolation on depression and anxiety. Despite some methodological limitations of these studies, there was fairly strong evidence that both social isolation and loneliness (perceived social isolation) were potentially detrimental in terms of development of mood disorders in later life.

The extant literature recommends that the effects of the individual components rather than a composite measure of social connection be used because the correlation between these components is relatively low. Investigating the effects of individual components could lead to greater insight into possible causal pathways and appropriate interventions. Epidemiological research has generally focused on the structural (e.g., social network size /density, marital status, living arrangements) or functional aspects of social relationships (received and perceived social support, perceived social isolation), while some researchers have also studied multi-dimensional approaches, i.e., a combination of structural and functional aspects. On the rare occasions where researchers have attempted to combine the effects of the components of social connection on the diagnostic outcomes, two different approaches have been used: 1. They have either used multivariable regression models (linear or logistic – depending on the outcome) with each of the components included as a separate predictor [34]. They have treated “social connection” as an underlying latent variable, while treating the individual components as the observed or “manifest” variables [65].

Inferences from this review should be drawn cautiously, considering some limitations. Firstly, we only included studies published in the English language. Other papers may be available in different languages. Secondly, we conducted a scoping review and not a systematic review; thus, the included papers were not assessed based on their quality. Some of the included studies in this review have small sample size. These studies must be replicated in larger, heterogeneous populations with a longer interval between the baseline assessment of social connectedness measures and follow-up of depressive and anxiety symptoms to ensure reliable and valid results for inferring the direction of causality. Despite these limitations, the longitudinal/prospective research reviewed here provides current knowledge on the effects of social connection on depression and anxiety and sheds important insights into possible causal mechanisms and a deeper understanding of how prior social circumstances can affect later mental health outcomes.

### Clinical and Policy Implications

The results of this scoping review have implications for clinical practice. Strong associations between lacking social connections and risk of depression underscore the importance of improving clinical detection of high-risk patients, including those with low received or perceived social support and elevated perceived loneliness. Because these characteristics may be difficult for primary care physicians to detect in their patients [66], consideration might be given to implementing brief screening instruments analogous to clinical tools that have been recommended to assess network size [67]. The link between social connections and improved adherence to medical recommendations [68] may motivate practitioners to detect and seek to address social isolation in their patients. However, meaningful clinical progress in addressing a lack of social connections may depend upon readily accessible and effective interventions.

The results also have implications for social policy. Because the prevalence of loneliness increases [69] and the size of social networks declines [70] as older adults age, the present findings bolster the public health rationale for addressing social vulnerabilities among older adults. Although some aspects of loneliness and social isolation likely involve genetic susceptibility [71], other aspects appear to involve characteristics of the neighborhood environment [72]. Policymakers seeking to reduce the adverse mental health effects of social isolation among older adults, for example, might weigh the benefits of designing supportive housing structures and neighborhoods for older adults that provide greater opportunities for socialization and formation of social connections.

## Data Availability

Yes - all data are fully available without restriction

---

**Preferred Reporting Items for Systematic reviews and Meta-Analyses extension for Scoping Reviews (PRISMA-ScR) Checklist**

| SECTION | ITEM | PRISMA-ScR CHECKLIST ITEM | REPORTED ON PAGE # |
| --- | --- | --- | --- |
| **TITLE** |  |  |  |
| Title | 1 | Identify the report as a scoping review. | 1 |
| **ABSTRACT** |  |  |  |
| Structured summary | 2 | Provide a structured summary that includes (as applicable): background, objectives, eligibility criteria, sources of evidence, charting methods, results, and conclusions that relate to the review questions and objectives. | 2 |
| **INTRODUCTION** |  |  |  |
| Rationale | 3 | Describe the rationale for the review in the context of what is already known. Explain why the review questions/objectives lend themselves to a scoping review approach. | 3 |
| Objectives | 4 | Provide an explicit statement of the questions and objectives being addressed with reference to their key elements (e.g., population or participants, concepts, and context) or other relevant key elements used to conceptualize the review questions and/or objectives. | 3 |
| **METHODS** |  |  |  |
| Protocol and registration | 5 | Indicate whether a review protocol exists; state if and where it can be accessed (e.g., a Web address); and if available, provide registration information, including the registration number. | N/A |
| Eligibility criteria | 6 | Specify characteristics of the sources of evidence used as eligibility criteria (e.g., years considered, language, and publication status), and provide a rationale. | 4 |
| Information sources* | 7 | Describe all information sources in the search (e.g., databases with dates of coverage and contact with authors to identify additional sources), as well as the date the most recent search was executed. | 4 |
| Search | 8 | Present the full electronic search strategy for at least 1 database, including any limits used, such that it could be repeated. | 4 |
| Selection of sources of evidence† | 9 | State the process for selecting sources of evidence (i.e., screening and eligibility) included in the scoping review. | 4 |
| Data charting process‡ | 10 | Describe the methods of charting data from the included sources of evidence (e.g., calibrated forms or forms that have been tested by the team before their use, and whether data charting was done independently or in duplicate) and any processes for obtaining and confirming data from investigators. | 4-5 |
| Data items | 11 | List and define all variables for which data were sought and any assumptions and simplifications made. | 4-5 |
| Critical appraisal of individual sources of evidence§ | 12 | If done, provide a rationale for conducting a critical appraisal of included sources of evidence; describe the methods used and how this information was used in any data synthesis (if appropriate). | N/A |
| Synthesis of results | 13 | Describe the methods of handling and summarizing the data that were charted. | 5 |
| **RESULTS** |  |  |  |
| Selection of sources of evidence | 14 | Give numbers of sources of evidence screened, assessed for eligibility, and included in the review, with reasons for exclusions at each stage, ideally using a flow diagram. | 5 & Fig1 |
| Characteristics of sources of evidence | 15 | For each source of evidence, present characteristics for which data were charted and provide the citations. | 5-13 |
| Critical appraisal within sources of evidence | 16 | If done, present data on critical appraisal of included sources of evidence (see item 12). | N/A |
| Results of individual sources of evidence | 17 | For each included source of evidence, present the relevant data that were charted that relate to the review questions and objectives. | 5-13 |
| Synthesis of results | 18 | Summarize and/or present the charting results as they relate to the review questions and objectives. | 5-13 |
| **DISCUSSION** |  |  |  |
| Summary of evidence | 19 | Summarize the main results (including an overview of concepts, themes, and types of evidence available), link to the review questions and objectives, and consider the relevance to key groups. | 13-14 |
| Limitations | 20 | Discuss the limitations of the scoping review process. | 15 |
| Conclusions | 21 | Provide a general interpretation of the results with respect to the review questions and objectives, as well as potential implications and/or next steps. | 15-16 |
| **FUNDING** |  |  |  |
| Funding | 22 | Describe sources of funding for the included sources of evidence, as well as sources of funding for the scoping review. Describe the role of the funders of the scoping review. | We did not include funding sources in the Acknowledgments or anywhere else in the manuscript file as is stated in the guidelines. Funding information was entered in the financial disclosure section of the submission system. |

JB1 = Joanna Briggs Institute; PRISMA-ScR = Preferred Reporting Items for Systematic reviews and Meta-Analyses extension for Scoping Reviews.
\* Where *sources of evidence* (see second footnote) are compiled from, such as bibliographic databases, social media platforms, and Web sites.
† A more inclusive/heterogeneous term used to account for the different types of evidence or data sources (e.g., quantitative and/or qualitative research, expert opinion, and policy documents) that may be eligible in a scoping review as opposed to only studies. This is not to be confused with *information sources* (see first footnote).
‡ The frameworks by Arksey and O'Malley (6) and Levac and colleagues (7) and the JBI guidance (4, 5) refer to the process of data extraction in a scoping review as data charting.
§ The process of systematically examining research evidence to assess its validity, results, and relevance before using it to inform a decision. This term is used for items 12 and 19 instead of "risk of bias" (which is more applicable to systematic reviews of interventions) to include and acknowledge the various sources of evidence that may be used in a scoping review (e.g., quantitative and/or qualitative research, expert opinion, and policy document).

## References

1. Becofsky KM, Shook RP, Sui X, Wilcox S, Lavie CJ, Blair SN. Influence of the source of social support and size of social network on all-cause mortality. Mayo Clin Proc. 2015;90(7):895–902.

2. Lamblin M, Murawski C, Whittle S, Fornito A. Social connectedness, mental health and the adolescent brain. Neurosci Biobehav Rev. 2017;80:57–68.

3. Ashida S, Heaney CA. Differential associations of social support and social connectedness with structural features of social networks and the health status of older adults. Journal of Aging and Health. 2008;20(7):872–93.

4. Holt-Lunstad J, Robles TF, Sbarra DA. Advancing social connection as a public health priority in the United States. Am Psychol. 2017;72(6):517–530.

5. Choi KW, Stein MB, Nishimi KM, Ge T, Coleman JRI, Chen CY, et al. An exposure-wide and mendelian randomization approach to identifying modifiable factors for the prevention of depression. Am J Psychiatry. 2020;177(10):944–954.

6. Silva M, Loureiro A, Cardoso G. Social determinants of mental health: A review of the evidence. The European Journal of Psychiatry. 2016;(4):259–92.

7. Ojagbemi A, Bello T, Gureje O. The roles of depression and social relationships in the onset and course of loneliness amongst Nigerian elders. Int J Geriatr Psychiatry. 2021;36(4):547–557.

8. Berthelsen M, Pallesen S, Magerøy N, Tyssen R, Bjorvatn B, Moen BE, et al. Effects of psychological and social factors in shiftwork on symptoms of anxiety and depression in nurses: A 1-year follow-up. Journal of Occupational and Environmental Medicine. 2015;57(10):1127–37.

9. Boyden JY, Hill DL, Carroll KW, Morrison WE, Miller VA, Feudtner C. The association of perceived social support with anxiety over time in parents of children with serious illnesses. Journal of Palliative Medicine. 2020;23(4):527–34.

10. Steine IM, Winje D, Krystal JH, Milde AM, Bjorvatn B, Nordhus IH, et al. Longitudinal relationships between perceived social support and symptom outcomes: Findings from a sample of adult survivors of childhood sexual abuse. Child Abuse & Neglect. 2020;107:104566.

11. Ciarleglio MM, Aslan M, Proctor SP, Concato J, Ko J, Kaiser AP, et al. Associations of stress exposures and social support with long-term mental health outcomes among US Iraq War Veterans. Behavior Therapy. 2018;49(5):653–67.

12. Porter E, Chambless DL. Social anxiety and social support in romantic relationships. Behavior Therapy. 2017;48(3):335–48.

13. Noteboom A, Beekman AT, Vogelzangs N, Penninx BW. Personality and social support as predictors of first and recurrent episodes of depression. Journal of Affective Disorders. 2016;190:156–61.

14. Van Den Brink RH, Schutter N, Hanssen DJ, Elzinga BM, Rabeling-Keus IM, Stek ML, et al. Prognostic significance of social network, social support and loneliness for course of major depressive disorder in adulthood and old age. Epidemiology and Psychiatric Sciences. 2018;27(3):266–277.

15. Aroian K, Uddin N, Blbas H. Longitudinal study of stress, social support, and depression in married Arab immigrant women. Health Care for Women International. 2017;38(2):100–17.

16. Hayslip Jr B, Blumenthal H, Garner A. Social support and grandparent caregiver health: One-year longitudinal findings for grandparents raising their grandchildren. Journals of Gerontology Series B: Psychological Sciences and Social Sciences. 2015;70(5):804–12.

17. Zhou M, Li F, Wang Y, Chen S, Wang K. Compensatory social networking site use, family support, and depression among college freshman: Three-wave panel study. J Med Internet Res. 2020;22(9):e18458.

18. Åhlin JK, Rajaleid K, Jansson-Fröjmark M, Westerlund H, Hanson LL. Job demands, control and social support as predictors of trajectories of depressive symptoms. Journal of Affective Disorders. 2018;235:535–43.

19. Billedo CJ, Kerkhof P, Finkenauer C, Ganzeboom H. Facebook and face-to-face: Examining the short-and long-term reciprocal effects of interactions, perceived social support, and depression among international students. Journal of Computer-Mediated Communication. 2019;24(2):73–89.

20. Scardera S, Perret LC, Ouellet-Morin I, Gariépy G, Juster RP, Boivin M, et al. Association of social support during adolescence with depression, anxiety, and suicidal ideation in young adults. JAMA Network Open. 2020;3(12):e2027491.

21. Crowe L, Butterworth P. The role of financial hardship, mastery and social support in the association between employment status and depression: Results from an Australian longitudinal cohort study. BMJ Open. 2016;6(5):e009834.

22. Houtjes W, Deeg D, van de Ven PM, van Meijel B, van Tilburg T, Beekman A. Is the naturalistic course of depression in older people related to received support over time? Results from a longitudinal population-based study. International Journal of Geriatric Psychiatry. 2017;32(6):657–63.

23. Misawa J, Kondo K. Social factors relating to depression among older people in Japan: analysis of longitudinal panel data from the AGES project. Aging & Mental Health. 2019;23(10):1423–32.

24. Stafford M, Antonucci TC, Zaninotto P. Joint trajectories of spousal social support and depressive symptoms in older age. Journal of Aging and Health. 2019;31(5):760–82.

25. Handley TE, Rich J, Lewin TJ, Kelly BJ. The predictors of depression in a longitudinal cohort of community dwelling rural adults in Australia. Social Psychiatry and Psychiatric Epidemiology. 2019;54(2):171–80.

26. Whitley DM, Kelley SJ, Lamis DA. Depression, social support, and mental health: A longitudinal mediation analysis in African American custodial grandmothers. The International Journal of Aging and Human Development. 2016;82(2-3):166–87.

27. Haverfield MC, Ilgen M, Schmidt E, Shelley A, Timko C. Social support networks and symptom severity among patients with co-occurring mental health and substance use disorders. Community Mental Health Journal. 2019;55(5):768–76.

28. Milgrom J, Hirshler Y, Reece J, Holt C, Gemmill AW. Social support—a protective factor for depressed perinatal women? International Journal of Environmental Research and Public Health. 2019;16(8):1426.

29. Nakamura A, Sutter-Dallay AL, El-Khoury Lesueur F, Thierry X, Gressier F, Melchior M, et al. Informal and formal social support during pregnancy and joint maternal and paternal postnatal depression: Data from the French representative ELFE cohort study. International Journal of Social Psychiatry. 2020;66(5):431–41.

30. Albuja AF, Lara MA, Navarrete L, Nieto L. Social support and postpartum depression revisited: the traditional female role as moderator among Mexican women. Sex Roles. 2017;77(3-4):209–20.

31. Asselmann E, Kunas SL, Wittchen HU, Martini J. Maternal personality, social support, and changes in depressive, anxiety, and stress symptoms during pregnancy and after delivery: A prospective-longitudinal study. Plos One. 2020;15(8):e0237609.

32. Asselmann E, Wittchen HU, Erler L, Martini J. Peripartum changes in social support among women with and without anxiety and depressive disorders prior to pregnancy: A prospective-longitudinal study. Archives of Women’s Mental Health. 2016;19(6):943–52.

33. Cankorur VS, Abas M, Berksun O, Stewart R. Social support and the incidence and persistence of depression between antenatal and postnatal examinations in Turkey: A cohort study. BMJ Open. 2015;5(4):e006456.

34. Chen HH, Chien LY. A comparative study of domestic decision-making power and social support as predictors of postpartum depressive and physical symptoms between immigrant and native-born women. PloS One. 2020;15(4):e0231340.

35. Faleschini S, Millar L, Rifas-Shiman SL, Skouteris H, Hivert MF, Oken E. Women’s perceived social support: associations with postpartum weight retention, health behaviors and depressive symptoms. BMC Women’s Health. 2019;19(1):1–8.

36. Hare MM, Kroll-Desrosiers A, Deligiannidis KM. Peripartum depression: Does risk versus diagnostic status impact mother–infant bonding and perceived social support? Depression and Anxiety. 2020;38(4):390–99.

37. Li Y, Long Z, Cao D, Cao F. Social support and depression across the perinatal period: a longitudinal study. Journal of Clinical Nursing. 2017;26(17-18):2776–83.

38. Racine N, Zumwalt K, McDonald S, Tough S, Madigan S. Perinatal depression: The role of maternal adverse childhood experiences and social support. Journal of Affective Disorders. 2020;263:576–81.

39. Razurel C, Kaiser B, Antonietti JP, Epiney M, Sellenet C. Relationship between perceived perinatal stress and depressive symptoms, anxiety, and parental self-efficacy in primiparous mothers and the role of social support. Women & Health. 2017;57(2):154–72.

40. Razurel C, Kaiser B. The role of satisfaction with social support on the psychological health of primiparous mothers in the perinatal period. Women & Health. 2015;55(2):167–86.

41. Senturk V, Abas M, Dewey M, Berksun O, Stewart R. Antenatal depressive symptoms as a predictor of deterioration in perceived social support across the perinatal period: A four-wave cohort study in Turkey. Psychological Medicine. 2017;47(4):766–75.

42. Tani F, Castagna V. Maternal social support, quality of birth experience, and post-partum depression in primiparous women. The Journal of Maternal-Fetal & Neonatal Medicine. 2017;30(6):689–92.

43. Zhong QY, Gelaye B, VanderWeele TJ, Sanchez SE, Williams MA. Causal model of the association of social support with antepartum depression: A marginal structural modeling approach. Am J Epidemiol. 2018;187(9):1871–1879.

44. Gan Y, Xiong R, Song J, Xiong X, Yu F, Gao W, et al. The effect of perceived social support during early pregnancy on depressive symptoms at 6 weeks postpartum: A prospective study. BMC Psychiatry. 2019;19(1):1–8.

45. Ohara M, Nakatochi M, Okada T, Aleksic B, Nakamura Y, Shiino T, et al. Impact of perceived rearing and social support on bonding failure and depression among mothers: A longitudinal study of pregnant women. Journal of Psychiatric Research. 2018;105:71–7.

46. Ohara M, Okada T, Aleksic B, Morikawa M, Kubota C, Nakamura Y, et al. Social support helps protect against perinatal bonding failure and depression among mothers: A prospective cohort study. Scientific Reports. 2017;7(1):1–8.

47. Chen HH, Hwang FM, Lin LJ, Han KC, Lin CL, Chien LY. Depression and social support trajectories during 1 year postpartum among marriage-based immigrant mothers in Taiwan. Archives Of Psychiatric Nursing. 2016;30(3):350–5.

48. Hetherington E, McDonald S, Williamson T, Tough S. Trajectories of social support in pregnancy and early postpartum: Findings from the All Our Families cohort. Social Psychiatry and Psychiatric Epidemiology. 2020;55(2):259–67.

49. Leonard KS, Evans MB, Kjerulff KH, Downs DS. Postpartum perceived stress explains the association between perceived social support and depressive symptoms. Women’s Health Issues. 2020;30(4):231–9.

50. Schwab-Reese LM, Schafer EJ, Ashida S. Associations of social support and stress with postpartum maternal mental health symptoms: Main effects, moderation, and mediation. Women & Health. 2017;57(6):723–40.

51. Tsai AC, Tomlinson M, Comulada WS, Rotheram-Borus MJ. Food insufficiency, depression, and the modifying role of social support: Evidence from a population-based, prospective cohort of pregnant women in peri-urban South Africa. Social Science & Medicine. 2016;151:69–77.

52. Yörük S, Açikgöz A, Türkmen H, Karlidere T. The prevalence of postpartum depression and the correlation of perceived social support and quality of life with postpartum depression: A longitudinal study. P R Health Sci J. 2020;39(4):327–335.

53. Zheng X, Morrell J, Watts K. Changes in maternal self-efficacy, postnatal depression symptoms and social support among Chinese primiparous women during the initial postpartum period: A longitudinal study. Midwifery. 2018;62:151–60.

54. Racine N, Plamondon A, Hentges R, Tough S, Madigan S. Dynamic and bidirectional associations between maternal stress, anxiety, and social support: The critical role of partner and family support. Journal of Affective Disorders. 2019;252:19–24

55. Morikawa M, Okada T, Ando M, Aleksic B, Kunimoto S, Nakamura Y, et al. Relationship between social support during pregnancy and postpartum depressive state: A prospective cohort study. Scientific Reports. 2015;5(1):1–9.

56. Domènech-Abella J, Mundó J, Haro JM, Rubio-Valera M. Anxiety, depression, loneliness and social network in the elderly: Longitudinal associations from The Irish Longitudinal Study on Ageing (TILDA). Journal of Affective Disorders. 2019;246:82–8.

57. Evans IE, Llewellyn DJ, Matthews FE, Woods RT, Brayne C, Clare L. Social isolation, cognitive reserve, and cognition in older people with depression and anxiety. Aging & Mental Health. 2019;23(12):1691–700.

58. Herbolsheimer F, Ungar N, Peter R. Why is social isolation among older adults associated with depressive symptoms? The mediating role of out-of-home physical activity. International Journal of Behavioral Medicine. 2018;25(6):649–57.

59. Holvast F, Burger H, de Waal MM, van Marwijk HW, Comijs HC, Verhaak PF. Loneliness is associated with poor prognosis in late-life depression: Longitudinal analysis of the Netherlands study of depression in older persons. Journal of Affective Disorders. 2015;185:1–7.

60. Chang SC, Pan A, Kawachi I, Okereke OI. Risk factors for late-life depression: A prospective cohort study among older women. Preventive Medicine. 2016;91:144–51.

61. Förster F, Stein J, Löbner M, Pabst A, Angermeyer MC, König HH, et al. Loss experiences in old age and their impact on the social network and depression–results of the Leipzig Longitudinal Study of the Aged (LEILA 75+). Journal of Affective Disorders. 2018;241:94–102.

62. Reynolds RM, Meng J, Dorrance Hall E. Multilayered social dynamics and depression among older adults: A 10-year cross-lagged analysis. Psychology and Aging. 2020;35(7):948–962.

63. Santini ZI, Jose PE, Cornwell EY, Koyanagi A, Nielsen L, Hinrichsen C, et al. Social disconnectedness, perceived isolation, and symptoms of depression and anxiety among older Americans (NSHAP): A longitudinal mediation analysis. The Lancet Public Health. 2020;5(1):e62–70.

64. Lund C, Brooke-Sumner C, Baingana F, Baron EC, Breuer E, Chandra P, et al. Social determinants of mental disorders and the sustainable development goals: A systematic review of reviews. Lancet Psychiatry. 2018;5(4):357–369.

65. Saeri AK, Cruwys T, Barlow FK, Stronge S, Sibley CG. Social connectedness improves public mental health: Investigating bidirectional relationships in the New Zealand attitudes and values survey. Australian & New Zealand Journal of Psychiatry. 2018;52(4):365–74.

66. Due TD, Sandholdt H, Siersma D, Waldroff FB. How well do general practitioners know their elderly patients’ social relations and feelings of loneliness? BMC Family Pract. 2018;19:34.

67. Pantell M, Rehkopf D, Jutte D, Syme SL, Balmes J, Adler N. Social isolation: a predictor of mortality comparable to traditional clinical risk factors. Am J Public Health. 2013;103(11):2056–62. DiMatteo MR. Social support and patient adherence to medical treatment: A meta-analysis. Health Psychol. 2004; 23(2):207–218.

68. Dykstra PA, van Tilburg TG, de Jong Gierveld J. Changes in older adult loneliness: Results from a seven-year longitudinal study. Res Aging. 2005; 27(6):725–747.

69. Wrzus C, Wagner J, Hänel M, Neyer FJ. Social network changes and life events across the life span: A meta-analysis. Psychol Bull. 2013; 139(1):53–80.

70. Goossens L, van Roekel E, Verhagen M, Cacioppo JT, Cacioppo S, Maes M, Boomsma DI. The genetics of loneliness: linking evolutionary theory to genome-wide genetics, epigenetics, and social science. Perspect Psychol Sci. 2015;10(2):213–26.

71. Tong S, Mullen RA, Hochheimer CJ, Sabo RT, Liaw WR, Nease De Jr, Krist AH, Frey JJ 3rd. Geographic Characteristics of Loneliness in Primary Care. Ann Fam Med. 2019;17(2):158–160.

